# OmiCrisp: A CRISPR SARS-CoV-2 test with Omicron detection

**DOI:** 10.1101/2023.01.06.23284282

**Authors:** Suruchi Sharma, Manasa Bagur Prakash, Nimisha Gupta, Vaijayanti Gupta, Vijay Chandru

## Abstract

We have developed a CRISPR based assay that can detect the presence of SARS-CoV-2 in RNA extracted from human samples and also predict if it is an Omicron or non-Omicron variant of the virus. This is a nucleic acid amplification-based test (NAAT). The amplification and detection are carried out in two independent steps in this assay. Amplification is done using a standard one-step RT-PCR method. The detection is done using a method that utilizes the trans-cleavage activity of the Cas12a enzyme. We have evaluated the performance of OmiCrisp in more than 80 clinical samples and observed an agreement of 100% with the sequencing results, in labeling SARS-CoV-2 positive samples as Omicron or non-Omicron. OmiCrisp -like platform can be developed quickly and can potentially complement sequencing for quick and rapid tracking of the transmission of new pathogen variants.

## Introduction

Regular testing for timely isolation has been a crucial activity in the management of the SARS-CoV-2 pandemic. Nucleic acid-based detection assays, real-time PCR (rtPCR), ubiquitously proved to be the most effective way of testing for the containment of Covid19 pandemic. This encouraged the development of new ways of nucleic acid-based tests that are cost-effective, rapid, and instrument agnostic (Vindeirinho et al., 2022). Furthermore, similar to all viruses, SARS-CoV-2 accumulates mutations in its genome over time. Some of these mutations may impact various properties of viruses including its infectivity, disease severity, response to therapy. Hence, in the absence of a complete knowledge of the severity of a given variant, it is of utmost important to track the transmission of the new SARS-CoV-2 variants for the effective management of the pandemic.

So far, for SARS-CoV-2 variant tracking, most countries have relied on whole genome sequencing of virus from patient samples. Indeed sequencing has been used to both understand the evolution of variants as well as track the variants by their signature mutations, once they have been sequenced (Vandenberg et al., 2021). Sequencing is expensive, time-consuming, and requires specialized instruments, and skilled personnel underscoring the need for a complementary rapid and cost-effective method for variant tracking. There are various cost-effective and rapid assays in published literature for genotyping of mutations (Little, 1995; Morlan et al., 2009). However, these are not being used in practice for tracking of SARS-CoV-2 variants because these methods require extensive design and validation efforts to develop, and hence not suitable for tracking of viruses which are evolving rapidly.

CRISPR systems appear to be a promising technology for sequence specific detection of nucleic acids. There have been various reports demonstrating CRISPR based methods for SARS-CoV-2 detection based on Cas13, Cas12, and Cas9 that show the promise of being instrument minimalistic (Azhar et al., 2021; Broughton et al., 2020; Ding et al., 2020; Fozouni et al., 2021; Guo et al., 2020; Joung et al., 2020; Kumar et al., 2021; Lucia et al., 2020; Nouri et al., 2021; Patchsung et al., 2020; Safari et al., 2021; Wang et al., 2021). Some of them have also shown variant detection capabilities, but most of these variant tracking platforms have not shown the variant selectivity at higher viral loads making them of limited use (Fasching et al., 2022; Kumar et al., 2021; Niu et al., 2022).

We here report the development OmiCrisp, OmiCrisp can detect the presence of SARS-CoV2 in an RNA sample extracted from a nasal or nasopharyngeal swab and predict if it is an “Omicron” or “non-Omicron” variant of the virus. This is a nucleic acid amplification-based test (NAAT). The amplification and detection are carried out in two independent steps in this assay. Amplification is done using a standard one-step RT-PCR method, followed by detection that relies on the trans-cleavage activity of the Cas12a enzyme (Chen et al., 2018; Li et al., 2018). Briefly, Cas12a enzyme binds to a guide RNA to make Cas12a: guide RNA complex. The guide RNA is customized to recognize the target DNA sequence -a sequence that one wants to detect. The Cas12a: guide RNA complex in the presence of target DNA makes a trimeric nucleoprotein complex, Cas12a: guide RNA: target DNA. This trimeric nucleoprotein complex has an endonuclease activity and it cleaves ssDNA irrespective of the sequence of ssDNA, this activity of the trimeric complex is known as trans-cleavage activity. The trans-cleavage activity of trimeric complex can be easily observed as increase in fluorescence signal if the ssDNA that gets cleaved is labeled with a fluorophore and quencher pair at its termini, reporter ssDNA. In our assay, the detection reagent contain Cas12a, a guide RNA, and ssDNA reporter, when a sample to be tested contains the target DNA to be detected by the guide RNA; the Cas12a: guide RNA: target DNA forms resulting in trans-cleavage of the ssDNA reporter and an increase in fluorescence signal. Total turnaround time of the assay is 3 hours. The assay has been designed to detect ORF1ab gene, N gene, and S gene targets for SARS-CoV-2; while ribonuclease P_MRP subunit (POP7) transcript target has been used as human RNA control.

OmiCrisp has been designed in such a way that the selectivity for variant prediction is retained at the highest possible viral load. Furthermore, we identified mutations in S gene and N gene which are very effective in discriminating Omicron from non-Omicron variants. These mutations are the basis for an agreement of 100% between OmiCrisp and sequencing results in calling a sample Omicron positive.

## Results and Discussion

### Guide design

In order to develop a nucleic acid-based assay for the detection of Omicron variant, we identified mutations that were specific to Omicron variant of SARS-CoV-2 and were not associated with any other SARS-CoV-2 variant identified by WHO; i.e. Alpha, Beta, Delta, Epsilon, Eta, Gamma, Iota, Kappa, Lambda, Mu, Theta, and Zeta. We enlisted mutations in the S gene of the Omicron variant that were present at a frequency higher than 90%, and not present in any of the aforementioned non-Omicron variants of SARS-CoV-2 at a frequency higher than 0.1 % (SI Table 1). Next, we selected the subset of these mutations that were present in all the three variants of Omicron, BA.1, BA.2, and BA.3, identified at that time of the work. This analysis was done manually from the data available at www.outbreak.info website on 6 January 2022.

In order to design a Cas12a trans-cleavage assay that can detect single nucleotide variations in a sequence, the variant should be present within the 6 bases of PAM site (Li et al., 2018). We examined the sequence of shortlisted mutation/s and identified the ones that matched the criteria for developing the Cas12a trans-cleavage-based assay for single nucleotide variation detection. We planned to develop an assay in which the variant prediction is based on the presence of a signal as opposed to absence of signal; hence, we designed a pair of guide RNAs to detect the sequences harboring each of these selected mutation/s. One of the guides of a pair is labeled as Reference-specific guide and another one is labeled as Omicron-specific guide. The Reference-specific guide RNA is designed to give a positive signal in trans-cleavage assay if the selected mutation/s is absent in the target, and the Omicron-specific guide RNA is designed to give a positive signal in a trans-cleavage if the selected mutation/s is present in the target.

In order to test the selectivity of the designed guide RNA pairs to discriminate Omicron from non-Omicron variants experimentally, the target region that harbors the mutation/s of interest, was amplified using one step RT-PCR with a suitable primer from synthetic RNA controls of indicated SARS-CoV-2 variant. The amplified product was used as target or input for the trans-cleavage assay. A guide pair qualified for being able to differentiate the Omicron from not Omicron targets, if a trans-cleavage assay done with the Omicron-specific guide showed at least 5 fold higher signal in presence of the target amplified from Omicron synthetic RNA control than in the presence of the target amplified from reference synthetic RNA control after 1 hour of the assay, and the trans-cleavage assay done using the “Reference-specific guide” showed at least 5 fold higher signal in presence of the target amplified from reference synthetic RNA control than in the presence of the target amplified from Omicron synthetic RNA control after 1 hour of the assay. Some of these designed guide pairs were indeed found to be able to differentiate Omicron from non-Omicron synthetic controls.

Our selective guides have shown the ability to differentiate Omicron from non-Omicron in end-point assays where we used ∼1*10^5^ copies of synthetic RNA control as input. However, at higher target concentrations the rates of trans-cleavage will be faster and the reactions for both Reference-specific and Omicron-specific guides may reach saturation after 1 hour of the assay, irrespective of the input RNA sample being Omicron or non-Omicron. Hence, it was important to evaluate if the selectivity of the guide RNA will be retained at higher input RNA concentrations in these end-point assay. The input to trans-cleavage assay is a DNA fragment amplified from the input RNA sample after RT-PCR amplification step. The maximum amount of the target DNA that can be expected at the end of RT-PCR amplification is equal to primer concentration used in the RT-PCR. In OmiCrisp, 250 nM of each primer is used for amplification; hence the highest possible concentration of the amplified product expected is 250 nM. 5 μL of the amplified product is employed in about 50 μL of reaction for trans-cleavage assay, so the highest possible concentration of the target DNA expected is 25 nM in the trans-cleavage reaction. So, it is reasonable to believe if a guide pair showed selectivity in the trans-cleavage assay performed in the presence of 30 nM of the synthetic DNA targets as inputs, the selectivity would be retained for any amount of viral RNA present in the unknown sample to be tested by the two-step assay.

Synthetic DNA templates corresponding to the amplicon region harboring the mutation/s of interest were synthesized as described in the materials and methods section. The trans-cleavage assays were done using these synthetic DNA targets (30 nM) as input for the selected guide pair.

The guide RNA pair was defined as suitable for the end-point assay if a trans-cleavage assay done with Omicron-specific guide showed at least 5 fold higher signal in the presence of Omicron synthetic DNA target (30nM) than in the presence of the reference synthetic DNA target (30 nM) after one hour of the assay, and trans-cleavage assay done with Reference-specific guide showed at least 5 fold higher signal in the presence of Reference synthetic DNA target (30 nM) than in the presence of the Omicron synthetic DNA (30 nM) after one hour of the assay. Two guide pairs that retained selectivity at high target DNA concentration: one pair targets a region on S gene and another targets the region on N gene (Figure 1b).

**Figure 1:**
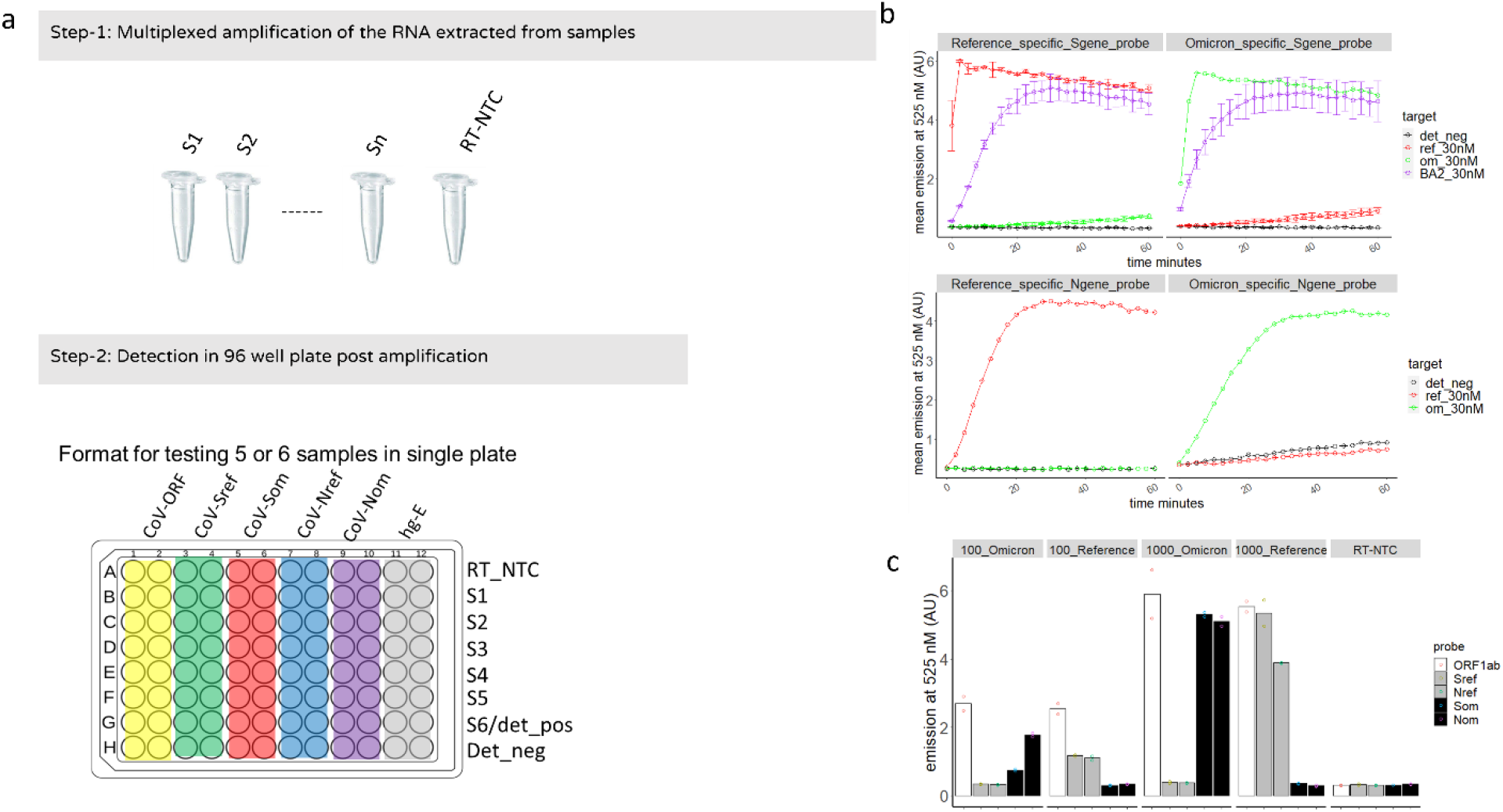
a) Two-step workflow of OmiCrisp. In step-1 or amplification step, S1, S2, up to S6 are RT-PCR reactions done with S1 to S6 to be tested. The RT_NTC is a control that is processed identical to the rest of the samples, except that instead of the sample, nuclease free water is added to it. Step-2 or the detection is done in a 96-well plate. In the schematic the wells of the plate are color coded to represent the guide RNA present in the well for detection. The sample that is added to each row is indicated at right. Det_pos indicates the mixture of synthetic DNA targets of all the six guides used in the assay, and det_neg indicates nuclease free water. RT-NTC, S1-to S6 are the amplified products obtained after the amplification step b) Trans-cleavage assay done at saturated concentration of the indicated DNA template with indicated guide RNA. This shows the selectivity of the guide pair for N and S gene to discriminate Omicron variant from non-Omicron variant is retained at saturating target DNA concentrations. c) The analytical validation of the OmiCrisp with specified synthetic RNA as input at indicated concentrations or input copies. The kinetics of the trans-cleavage reaction was followed over time with high synthetic template concentrations of 30nM. Specificity of the guides for the respective templates is indicated. The sensitivity of the assay is established at 100 and 1000 copies of synthetic template for an end-point assay readout at 60 minutes. Note, OmiCrisp_v1 did not have Nom and Nref guide RNAs containing detection reagents, hence only detection was done in presence of only four different detection reagents for it.

The S gene guide pair that we selected targets a region on S gene that harbors a set of three mutations; Q493R, G496S, and Q498R. The BA.2 variant of Omicron does not harbor the G496S mutation. Interestingly, we observed that both the reference-specific and Omicron-specific guide of this guide pair can induce trans-cleavage in the presence of the synthetic DNA template corresponding to BA.2. So, the samples that will show trans-cleavage in the presence of both Omicron- and Reference specific guide will still be Omicron. We will have to keep testing for all the evolving variants what the results will be if they do not harbor all three mutation/s. See, methods sections for the detailed calculations for the variant prediction of the assay. The guide pair for the N gene that we selected targets a region on the N gene that harbors mutation NΔ31-33 (figure 1b).

In the first version of OmiCrisp, OmiCrisp_v1, we did not include the N gene guide pair for detection. As described ahead, upon completion of the validation of OmiCrisp_v1 we observed that S gene gives lower or undetectable signals at low sample loads, see figure 2, hence compromising the sensitivity of assay of SARS-CoV-2 prediction. To overcome this limitation, we included a guide pair for the N gene in our version 2 of the OmiCrisp (Figure 1c).

**Figure 2:**
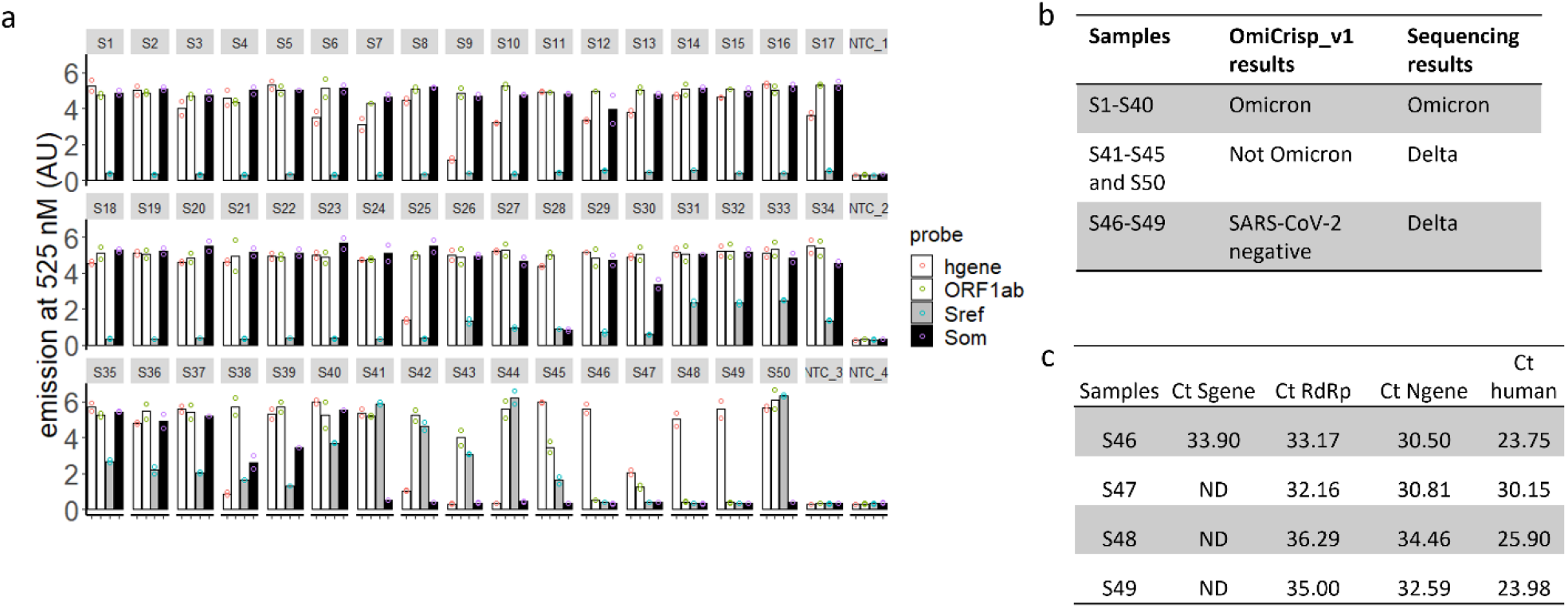
Clinical validation of OmiCrisp_v1. a) Bar graph showing the fluorescence intensity of the indicated probe or guide RNA for each sample. The height of the bar represents the background subtracted mean intensity of the technical duplicates in arbitrary fluorescence units (AU), and the empty circle overlaid on bars indicate the background subtracted intensity of the individual technical duplicates. b) Tabular summary of the clinical validation, showing the comparison between sequencing and Omcrisp_v1. c) qRT-PCR analysis of the sample identified as SARS-CoV-2 negative by the OmiCrisp_v1.

### Primer design

We designed the primers that could amplify the regions of interest, harboring the target of the guide RNA to be used for detection, of S, ORF1ab, and N genes. In order to ensure that these primers will be applicable for the majority of SARS-CoV-2 variants, we designed consensus primers for a group of 23 variants of SARS-CoV-2 that represent measure mutation constellations of SARS-CoV-2, using NCBI primer BLAST (SI table 2 for the variants selected for primer design). In order to estimate the applicability of these primers for Indian isolates of SARS-CoV-2, we aligned these primers with all the complete genome sequences of SARS-CoV-2 Indian isolates deposited at NCBI. We estimated the frequency of isolates that have mis-matches with primer. We observed that 99 % of these isolates have no mis-matches with the designed primers. The sequence of primers and the frequency of isolated with mis-matches for each prime is provided in SI table 3. Also, the N gene primer was modified to insert a PAM site in the amplicon because the mutation/s of interest in the N gene did not have a PAM site closer than 6 bases from it.

### Assay validation

Figure 1a shows the workflow of the two-step assay. Step-1 or the multiplexed amplification was carried out in PCR tubes. RT-NTC control is included at this step, in this control nuclease free water is added to the indicated tube instead of a sample. Step-2 or detection step was carried out in a 96-well plate. Briefly, a fragment of interest of ORF1ab, N, S, and human RNaseP gene was amplified from the sample to be tested using about 5 μL of the sample to be tested in 70 μL of the reaction volume using multiplexed one step RT PCR as described in materials and methods section. In the second step, 5 μL of the amplified product was used as an input for end-point trans-cleavage assay in the presence of 6 different detection reagents containing the following guide RNAs: 1. ORF1ab (ORF1ab_ref_ + ORF1ab_om_) 2. S_om_ 3. S_ref_ 4. N_ref_ 5. N_om__om and 6. RNAseP (SI table 5 for guide RNA sequences). Trans-cleavage reaction in the presence of each guide RNA was done in duplicate, resulting in a total of 12 independent trans-cleavage reactions for each sample to be tested. OmiCrisp_v1 did not have N_om_ and N_ref_ guide, hence detection was done in presence of only four different detection reagents leading to total 8 independent trans-cleavage for each sample.

### Analytical validation

For analytical validation a known number of copies of a given synthetic RNA control was used as a sample to be assayed. As can be seen in figure 1c when 100 copies of either Omicron or Reference synthetic RNA control samples were used as input to the assay a detectable signal higher than RT-NTC was observed at the end of the assay; hence the limit of detection of the assay was expected to be at least 100 copies of RNA. It was observed that the Reference synthetic controls samples showed more than 2 times higher signal in the trans-cleavage reactions that had Reference-specific guides (S_ref_ or N_ref_) than in the trans-cleavage reactions that has Omicron-specific guide of corresponding gene (S_om_ or N_om_), and Omicron synthetic controls samples showed more than 2 times higher signal in the trans-cleavage reactions that had Omicron-specific guides (S_om_ or N_om_) than in the trans-cleavage reactions that had Reference-specific guide of corresponding gene (S_ref_ or N_ref_). Hence, the N gene and S gene guide pair were able to discriminate the Omicron synthetic RNA from reference synthetic RNA at all the concentration of input RNA tested using the assay.

### Clinical Validation

We validated the OmiCrips_v1 in RNA samples extracted for form nasopharyngeal swabs that were previously sequenced and identified as SARS-CoV-2 positive. These blinded identity samples were provided by Strand Life Sciences, an accredited NGS genomic surveillance center in Bengaluru, India. As mentioned previously, the first version of OmiCrisp, OmiCrisp_v1, comprised four guide RNAs. Three of them are for SARS-CoV-2: ORF1ab, S gene Omicron-specific, S gene reference-specific, and one for human gene: RNase P gene. For clinical validation 5 μL of extracted RNA from a clinical sample was used as an input to the assay.

Out of the 50 samples tested only 46 were detected as SARS-CoV-2 positive by OmiCrisp_v1; therefore, the sensitivity of the assay for detecting SARS-CoV-2 was 92 %. Among the 46 samples detected as SARS-CoV-2 positive by OmiCrisp_v1, 40 were identified as Omicron and 6 were identified as non-Omicron. All the samples identified as Omicron by OmiCrisp_v1 were indeed Omicron, the sample identified as “non-Omicron” were Delta (non-Omicron) as per sequencing results. Therefore, the specificity of OmiCrisp_v1 for distinguishing Omicron from non-Omicron variants is 100 % (Figure 2a, 2b).

OmiCrisp_v1 had a good selectivity to call out Omicron from non-Omicron variants; however, in order to improve the sensitivity of calling SARS-CoV-2 positive, we closely looked at the data of 4 samples that were identified as SARS-CoV-2 negative. We observed that two of these samples have unambiguously higher signals over no template controls for ORF1ab gene. But because of the S gene signal was low, these samples were identified as negative (Figure 2a). Next, we estimated the Ct values of all the four samples using a commercially available rtPCR kit that targets S gene, N gene, and RdRP gene (Figure 2c). Interestingly in three out of these four samples S gene was not detectable, and the Ct values of the N-gene, and RdRp were higher than 32 in these samples. This indicates lower viral load or degraded RNA in these samples.

Based on the above data, to enable detection at viral lower loads, we introduced two changes to OmiCrisps_v1 to build the improved OmiCrisp_v2. First, we increased the number of PCR cycles from 35 to 40 at the amplification step, and we introduced a new SARS-CoV-2 gene N gene to the assay.

We validated the Omcrisp_v2 on a total of 33 RNA samples extracted from nasopharyngeal swabs. This validation was done at inStem and conducted by the inStem technicians (Bengaluru, India) in a blinded fashion. The unprocessed data with blinded sample identities was handed over to CrispBits by inStem (Figure 3c). CrisprBits analyzed the data and made predictions based on OmiCrisp_v2 (Figure 3b). We developed an automated pipeline for the data analyses of OmiCrisp assay that is independent of technician’s subjective error. The details are in the method section.

**Figure 3:**
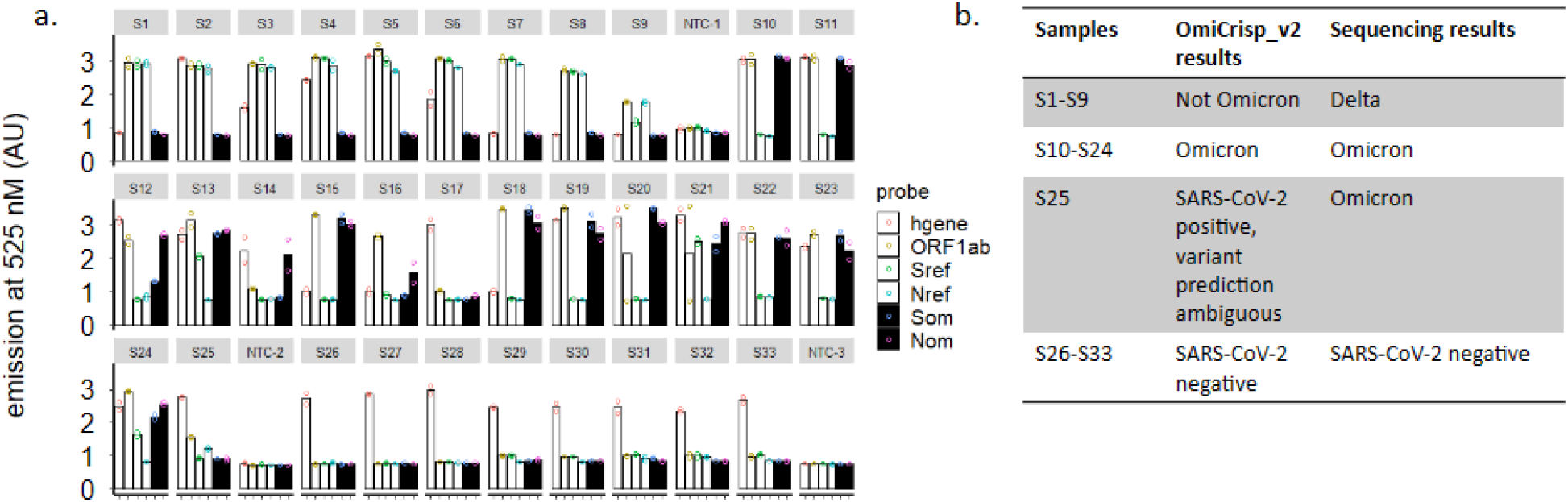
Clinical validation of OmiCrisp_v2. a) Bar graph showing the average fluorescence intensity of the indicated probe or guide RNA for each sample. The height of the bar represents the mean intensity (arbitrary fluorescence unit (AU) of the technical duplicates, and the empty circle overlaid on bars indicate the intensity of the individual technical duplicate. b) Tabular summary of the clinical validation, showing the comparison between sequencing and OmiCrisp_v2. *\*validation was performed in an external laboratory where the qPCR instrument was used to measure end point fluorescence of the CRISPR assay*

Out of 33 samples 25 samples were SARS-CoV-2 positive and 8 samples were SARS-negative. OmiCrisp_v2 correctly identified the positive sample as SARS-CoV-2 positives and negative samples as SARS-CoV-2 negative. Hence, the specificity and sensitivity of OmiCrisp_v2 assay in identifying SARS-CoV2 in this validation study were both at 100 %.

Out of 25 positive samples, 9 samples were Delta variants of the SARS-CoV-2 virus and OmiCrisp_v2 correctly identified them as non-omicron. Among the remaining 16 samples that were Omicron 15 were accurately identified as Omicron. One of the Omicron samples was labeled as ambiguous by OmiCrisp_v2 because it was identified as Omicron with S gene guide pair and as non-Omicron as N gene guide pair.

The validation results indicate that OmiCrisp platform is suitable for tracking the Omicron variant of SARS-CoV-2 variant in clinical samples.

### Conclusions

We developed a CRISPR based assay, OmiCrisp, that can predict the presence of Omicron or non-Omicron variant in extracted RNA samples. We validate the results of the assay on RNA samples extracted from human nasopharyngeal swabs. We observed that OmiCrisp is highly reliable in identifying the variants. The assay interpretation is streamlined and can be easily automated, and does not require any subjective decisions to be made by technicians performing the assay. The clinical validation of the assay was performed at an external laboratory indicating that the assay is easily amenable to technology transfer to any lab.

Kumar et al have also reported the development of a CRISPR systems-based platform for SARS-CoV-2 variant detection that is based on using FnCas9, named RAY (Kumar et al., 2021). This is an endpoint assay, and in end-point assay the selectivity can be lost if the concentration of input RNA is high. However, the validation for the study was performed on samples with Ct values above 22, indicating higher concentration of target DNA in these samples. The Ct values in samples from SARS-CoV-2 positive patients for ∼20-50 % of samples is below 22 (Buchan et al., 2020; Platten et al., 2021). We have designed OmiCrisp to ensure the variant selectivity will not be compromised at very high viral loads (Figure 1b).

Another interesting CRISPR based platform for variant detection is miSHERLOCK which is based on Cas13a, where the input is saliva samples from prospective patients and nucleic acid extraction is done automatically in the device that is included in the platform. However, the platform performance has not been shown in clinical samples, and the variant calling specificity has not been tested at higher concentrations of RNA (de Puig et al., 2021).

Fasching et al identified a novel Cas12a enzyme, CasDx1, for improved SNP detection and showed its application in SARS-CoV-2 variant detection (Fasching et al., 2022). Again, there is no explicit experiment evidence showing the selectivity of guide RNAs at higher concentrations on the input samples. The agreement of sequencing and the assay for the variant identification varies from 83% (Alpha) - 97 % (WT).

OmiCrisp is a CRISPR based assay that can be used for the detection of SARS-CoV-2 Omicron variants. OmiCrisp depends on the presence of a positive signal to call out a sample either Omicron or non-Omicron that ensures that the assay is more reliable for variant prediction even in samples with lower viral load. An assay that labels a sample as Omicron or non-Omicron based on absence of signal from a given gene will have very low specificity at lower viral loads. For example, an rtPCR that identifies non-Omicron based on absence of S gene signal would have an identified sample 46, 47, 48 used in OmiCrisp_v1 validation as Omicron whereas in reality they were non-Omicron (figure 2c) (Thermo Fisher, 2021).

The design, development and validation of a nucleic acid-based assay, like OmiCrisp, for variant detection has rapid turnaround time and relatively lower investment costs in the R&D. Hence, these types of assays can complement ongoing sequencing efforts in a facility for quick screening of samples before selecting them for sequencing or to validate suspicious sequencing results. Furthermore, the OmiCrisp assay can be done with relatively lower quality and quantity of nucleic acid samples than that required for NGS. This makes OmiCrisp and similar CRISPR based assays a choice for screening of variants in relatively poor-quality samples that fail in NGS. In summary, Omi Crisp is an example of an accurate, rapid and cost-effective platform that can be helpful in tracking fast evolving pathogens.

From the onset, environmental samples, specifically wastewater samples, have shown great promise in getting the early warning of the upcoming peak (Zhu et al., 2021). Furthermore, waste water surveillance is not dependent on access to healthcare and testing capacity of clinical labs (Kirby et al., 2021). However, an environmental sample exposed to various conditions that can degrade nucleic acids are not expected to yield very high quality RNA. OmiCrisp platform does not require highly pure or concentrated samples as input. We postulate that OmiCrisp and similar CRISPR assays can be utilized for monitoring of pathogens and their variants in the environment samples for variant tracking, and thus extending their use to pandemic surveillance.

## Materials and method

### Reagents and Equipment

LbaCas12a, Alt-R™ Lb. Cas12a (Cpf1) Ultra, custom guide RNAs Alt-R® Lb. Cas12a crRNA, custom ssDNA_FQ reporter (/56-FAM/TT ATT /3IABkFQ/) were procured from IDT(Integrated DNA Technologies, USA). DNA oligonucleotides used as primers and synthetic templates were custom synthesized from Sigma (USA), and VNIR Biotechnologies (India). NEBuffer™ 2, and BSA (B9000S) were purchased from NEB(New England BioLabs, USA). The fluorescence measurements were done in VarioSkan LUX micro plate reader (Thermo Scientific, USA) in Corning® 96 Well Black Polystyrene Microplates (CLS3603-48EA, Sigma, USA) were used. The fluorescence measurements for OmiCrisp_v2 were acquired in Bio-Rad CFX96 Real-Time PCR machine (Bio-rad, USA). The amplification reactions were performed in Mastercycler Nexus Thermal Cycler™ (Eppendorf, Germany) and Applied Biosystems 2720 thermal cycler (Thermo Scientific, USA). SARS-CoV-2 synthetic RNA controls for reference strain (Wuhan-Hu-1, cat. 102024) and Omicron strain BA.1 (Omicron EPI_ISL_6841980, cat.105204) were procured from TwistBioscieces, USA. PCR master mix 2X, K01721, was procured from Thermo Scientific. For purification of PCR products QIAquick Gel Extraction Kit (cat.28704, Qiagen, Germany) was used.

### Synthetic DNA template preparation

Synthetic DNA templates corresponding to S gene Omicron BA.1, and S gene Omicron BA.2 were prepared by the overlap extension of the synthetic oligonucleotides. Briefly, the overlap extension mix was prepared by mixing overlapping oligonucleotides, see SI table 4 for sequences, at a concentration of 1 μM in 1X PCR master mix and overlap extension was performed in thermal cyclers with following conditions: Initial heat denaturation 95 °C for 5 minutes; 15 cycles of 90 °C 30 seconds heat denaturation, annealing at 60 °C for 30 seconds, extension 72 °C for 30 seconds; and final extension for 5 minutes at 72 °C. Rest of the synthetic templates were prepared by One step RT-PCR amplification from the corresponding synthetic RNA controls with the suitable primers at 250 nM concentration and 1*10^5 copies of synthetic RNA as template. The products obtained after overlap extension and one step RT-PCR were purified using a purification kit and quantified based on absorbance at 260 nm in NanoDrop One™ (Thermo Scientific, USA).

### One step RT-PCR

One step RT PCR was performed using commercially available one step RT PCR mix. The concentration of each primer in the reaction mix was about 250 nM. Following thermal cycling conditions were used for the one step RT-PCR: Step 1. Primer incubation at about 25 °C for about 2 minutes, Step 2. Reverse transcription at about 53 °C for about 10 minutes, Step 3. Enzyme denaturation at about 95 °C for about 2 minutes, and Step 4. 35-40 cycles of amplification that includes incubation at about 95 °C for about 15 seconds for denaturation and combined extension/annealing at about 60 °C for about 1 minute. For OmiCrisp_v1 and OmiCrisp_v2 multiplexed one step RT PCR was done. OmiCrisp_v2 used 4 primer pairs to amplify fragments of interest corresponding to ORF1ab, S gene, N gene, and human RNaseP; hence a total of 8 primers were added in a single tube. For OmiCrisp_v1 N gene primers were not included.

### Trans-cleavage assay

All the trans-cleavage assays for a given target template or sample were carried out using 25 nM of LbaCas12a, 25 nM of the indicated guide RNA, 50 nM ssDNA_FQ reporter in a solution containing 50 mM NaCl, 10 mM Tris-HCl, 10 mM MgCl_2_, and 100 μg/ml BSA pH 7.9 (at 25 °C) at 37 °C. For the end-point assays the reactions were stopped after 1 hour of incubation by adding 10 μL of stop buffer, 250 mM EDTA and 37.5 mM Tris-HCl pH 7 at 25 °C to the 50 μL reaction mix. For real time assays, all the reactions were initiated by addition of the target samples, and it was ensured that the time difference between beginning of addition of sample and the start of the data acquisition is not more than 2 minutes. Fluorescence data (emission at 525 nM) was captured using either a plate reader or real time PCR machine. Data is shown in an arbitrary fluorescence unit (AU).

### Clinical samples for validation

Clinical nasopharyngeal swab samples were obtained from two sources. (1) Strand Life Sciences which was mandated by the Technical Advisory Committee (TAC) of the State of Karnataka for Pandemic to initiate the genomic Surveillance of SARS-CoV-2 in various parts of Bengaluru city (Ref: CHO (PH)/PR/P-103/2021-22), helped us to cross validate OmiCrisp v1 of the assay. Ethics for sample collection and research was provided by the institutional ethics committee at HCG (Health Care Global), and (2) The Institutional Ethics Committee, inStem, provided the approval for archival and access of clinical samples from the biorepository at inStem (inStem/IEC-19/01/27E, inStem/IEC-17/001). The Institutional Biosafety Committee provided permission (inStem/G-141(3)-17/CJ) for the utilization of the samples for clinical research. Permission to conduct and support research on SARS-CoV2 has been provided to inStem by the Review Committee on Genetic Manipulation (RCGM), Department of Biotechnology, Ministry of Science and Technology, Government of India (approval number: BT/IBKP/035/2019).

### Data analysis and interpretation

#### Threshold signal estimation

The threshold signal is important for labeling a given signal as unambiguously positive without requiring a subjective decision of the analyst. The threshold signal was calculated as described below. Every time a trans-cleavage assay was performed, negative controls were included for each guide RNA used for the detection in trans-cleavage assay. The negative controls had all the reagents except the sample added to them. Signals from negative controls were used to estimate the threshold signal. The signal from the negative controls of all the guide RNAs are pooled into one set that was labelled as “pooled negative controls”. The median and interquartile range of the “pooled negative controls” was calculated using standard formulas. First, outliers, which could arise from random contamination, were removed from the “pooled negative controls”. Upper limit of outliers was calculated using equation **Eq.1**.

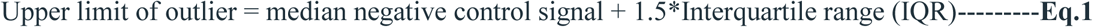

All the negative control points with signals higher than the Upper limit of outlier were removed. In case of a situation where more than 20 % of the negative controls were higher than the upper limit of outlier, the entire assay was discarded and repeated with fresh reagents. After removal of outliers, standard deviation was calculated of the remaining “pooled negative data set” using standard formulas and Threshold signal (Thresh_signal) was calculated using equation **Eq.2**.

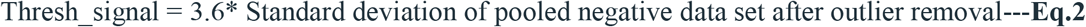

#### RT-NTC check

The RT-NTC control is the control where no sample was added at the step of amplification to the RT-PCR reaction. The presence of signal higher than threshold signal in the RT-NTC for any guide RNA indicated contamination. A criterion for the acceptable limit of contamination, RT-NTC cut off, was set. If for any guide RNA the signal in RT-NTC was more than RT-NTC cut off the data for the guide RNA was not used for analysis. The RT-NTC cut off was calculated using equation **Eq.3**.

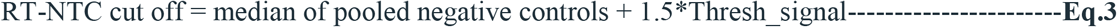

#### Unreliable data removal

In order to interpret results of the assay for a given sample, first the reliability of the signals obtained after trans-cleavage reaction with all guide RNAs was evaluated. The reliability of the signals was estimated by estimating the variations in technical duplicates of the trans-cleavage assay for each guide for each sample. And any data points which showed large variations in duplicates were considered unreliable and were excluded from the data analysis. In order to remove the data points with large variation in signal in technical duplicates, percentage relative standard deviation (%RSD) for technical duplicates was calculated using equation **Eq.4**.

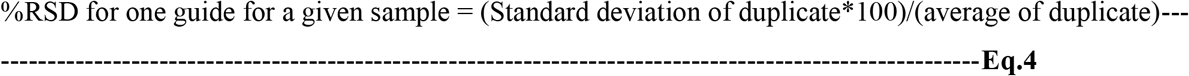

All the data points with %RSD more than 20% were removed from the analysis. The data points with human gene guide RNA %RSD more than 20% were not removed from the analysis. In cases where after removing the unreliable data, the data for less than three SARS-CoV-2 guides was left, the assay for that sample was repeated. Note, the exception was applied to two samples used for OmiCrisp_v1 validation. The S om guide of sample 12 and ORF1ab guide of S40 had %RSD of 28 and 20 %, respectively. We did not have sufficient sample for the repeat; however, the signal of the duplicate was approximately 10 times higher than the background, so it was reasonable to believe that these signals were not unreliable.

#### Labeling the signal as positive or negative

In order to label a signal as positive or negative, the average signal of all the technical duplicates left after the unreliable data removal was estimated. Next, the background subtracted signal was estimated for each guide RNA used in the assay for each sample tested. The background subtracted signal (back_sub_signal) for all these average signals was calculated using equation Eq.5.

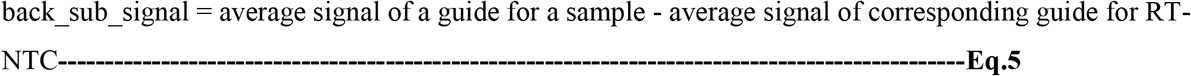

If the background subtracted signal was greater than threshold signal it was considered positive signal else it was considered negative.

#### SARS-CoV-2 positive versus negative prediction

1. The samples that did not positive signal for any SARS-CoV-2 genes were labeled as “SARS-CoV-2 negative”.
2. The samples that showed positive signal for only one SARS-CoV-2 gene (out of ORF, S, and N) were labeled as “SARS-CoV-2 negative” with a remark saying, “one gene positive repeat to confirm”.
3. The samples that showed positive signal for two or threeSARS-CoV-2 genes (out of ORF, S, and N) were labelled as “SARS-CoV-2 positive”.

Notably, if either one or both guides of a guide pair for a gene showed a positive signal it was counted as only 1 gene positive. For example, the S gene had two guides: S_ref_ and S_om_. S gene was counted as one gene positive if either one or both the S gene guides showed positive signal. Similarly, the N gene has two guides: N_ref_ and N_om_. The N gene was counted as one gene positive if either one or both the N gene guide RNA showed a positive signal.

### Omicron versus non-Omicron prediction

#### Variant prediction using S gene guide RNA pair

Based on the signals observed for S gene guide pair, S_om_ and S_ref_, following two situations were possible.

1. For samples where the back_sub_signal for both S_om_ and S_ref_ signals were not available, variant prediction was not made with S gene saying “insufficient data for prediction of variant”.
2. For samples where both S_om_ and S_ref_ back_sub_signal were available, S_om_/S_ref_ was calculated using equation **Eq.6**. If, So_m_/S_ref_ >0.5, the sample was labeled as “Omicron”, else it is labeled as “Not-Omicron”.

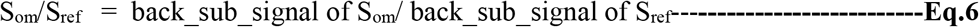

#### Variant prediction using N gene guide pair

Based on the signals observed for N gene guide pair, N_om_ and N_ref_, following two situations were possible.

Based on the signals observed for N gene guide pair, N_om_ and N_ref_, following two situations were possible.

1. For samples where the back_sub_signal for both N_om_ and N_ref_ signals were not available, variant prediction was not made with N gene saying “insufficient data for prediction of variant”.
2. For samples where the back_sub_signal for both N_om_ and N_ref_ were available, N_om_/N_ref was_ calculated using equation Eq.7. If, N_om_/N_ref_ >1, the sample was labelled as “Omicron”, else it was labelled as “Not-Omicron”.

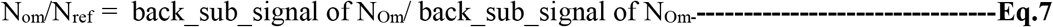

#### Final variant prediction was done as described below

1. If only one of the N or S gene could be used for variant prediction, the results corresponding to the variant prediction of that gene was reported.
2. If both N and S gene could be used for prediction and the prediction was same for both the corresponding result was reported.
3. If N and S gene variant predictions do not match the result reported was, “N and S gene predictions do not match, possible contamination or mixed samples repeat with freshly collected samples.”
4. If neither N nor S gene could be used for variant prediction the result was, “assay needs to be repeated because the data was not sufficient for variant prediction.”

#### Human gene signal analysis

If the signal for human gene was positive the result reported was “human gene detected” else the result reported was “human gene not detected”. If the sample SARS-CoV-2 negative and human gene was also not detected the remark was added assay needs to be repeated with fresh sample or higher amount of sample.

## Data Availability

All data produced in the present study are available upon reasonable request to the authors and also included in the manuscript

## Acknowledgements

We acknowledge the C-CAMP-InDx Program (Bengaluru, India) for providing us resources and technical support for analytical and clinical validation of the OmiCrisp assay. The validation of the OmiCrisp has been conducted with the significant contribution and expertise of the DBT-inStem biorepository and the COVID testing laboratory (Bengaluru, India). STRAND life sciences (Bengaluru, India) provided validation support for the work. We acknowledge Mr. Lalith Kishore, CEO, InDx, Dr. Chitra Pattabiraman and Dr. Reety Arora for their scientific inputs and discussions.

## SI tables

**SI table 1:** The table shows the list of manually selected in S gene of SARS-CoV-2 that are selective for the Omicron variant.

| Omicron specific S gene mutations from outbreak.info on 6-01-2022 | Notes |
| --- | --- |
| N211I del212 | The frequency was below 90% |
| G339D | Selected for guide design |
| S371L, S373P, S375F | No PAM site |
| E484A | No PAM site |
| Q493R/G496S/Q498R | Selected for guide design |
| T547K | Not present in BA.2 |
| N679K | Selected for guide design |
| N856K | Selected for guide design |
| Q954H | No PAM site |
| N969K | Selected for guide design |
| L981F | Not present in BA.2 |

**SI table 2:** The table shows the list of variants selected for designing the consensus primers for SARS-CoV-2. The consensus sequence for each variant was obtained by aligning 200 sequences of corresponding lineage available at NCBI. In cases where 200 sequences were not available all the available sequences of the variant were used for the obtaining the consensus sequence.

| S. No. | The Pango lineages representing WHO identified variant constellations | Number of complete sequences in NCBI |
| --- | --- | --- |
| 1 | A.23.1 | 203 |
| 2 | AV.1 | 100 |
| 3 | AY.4.2.1 | 531 |
| 4 | AY.4 | 40968 |
| 5 | B.1.1.318 | 16 |
| 6 | B.1.1.7 | 195057 |
| 7 | B.1.351 | 723 |
| 8 | B.1.427 | 4144 |
| 9 | B.1.429 | 9493 |
| 10 | B.1.525 | 722 |
| 11 | B.1.526 | 19274 |
| 12 | B.1.617.1 | 221 |
| 13 | B.1.617.2 | 10249 |
| 14 | B.1.617.3 | 1 |
| 15 | B.1.621 | 29 |
| 16 | BA.1 | 73446 |
| 17 | BA.2 | 2974 |
| 18 | BA.1.1 | 142638 |
| 19 | C.37 | 409 |
| 20 | P.1 | 6381 |
| 21 | P.2 | 504 |
| 22 | P.3 | 12 |
| 23 | AY.122 | 4127 |
| 24 | B.1.247 | 38 |
| 25 | B.1.1.216 | 50 |
| 26 | B1.1.36 | 1017 |
| 27 | A.7 | 4 |
| 28 | B.1.36.8 | 285 |
| 29 | B.1.1 | 9189 |

**SI table 3:**
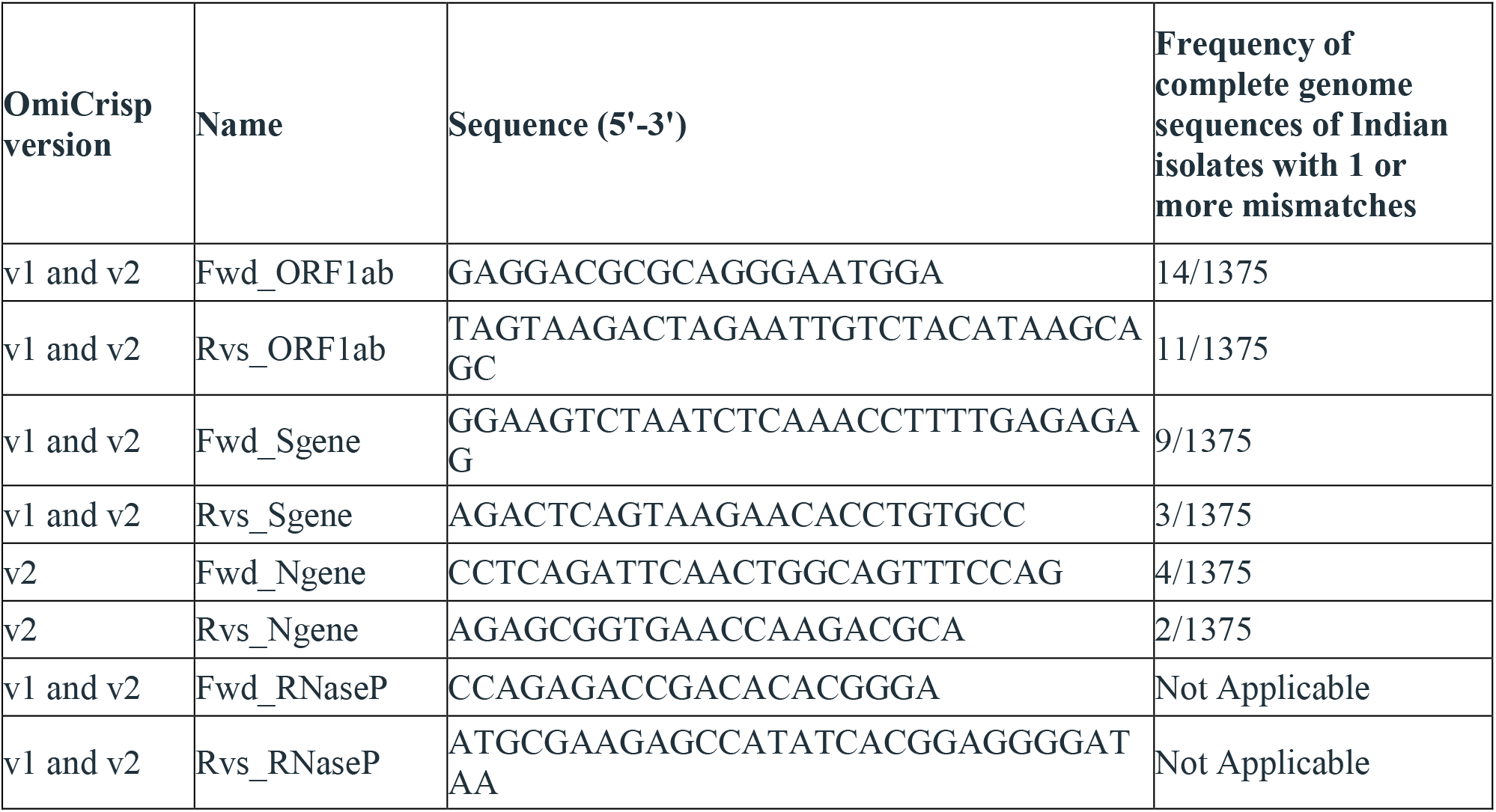
The list of primers used in multiplexed RT-PCR of OmiCrips_v1 and OmiCrisp_v2. Frequency of mismatches in the primers with the Indians isolates have also been shown in the table.

**SI table 4:**
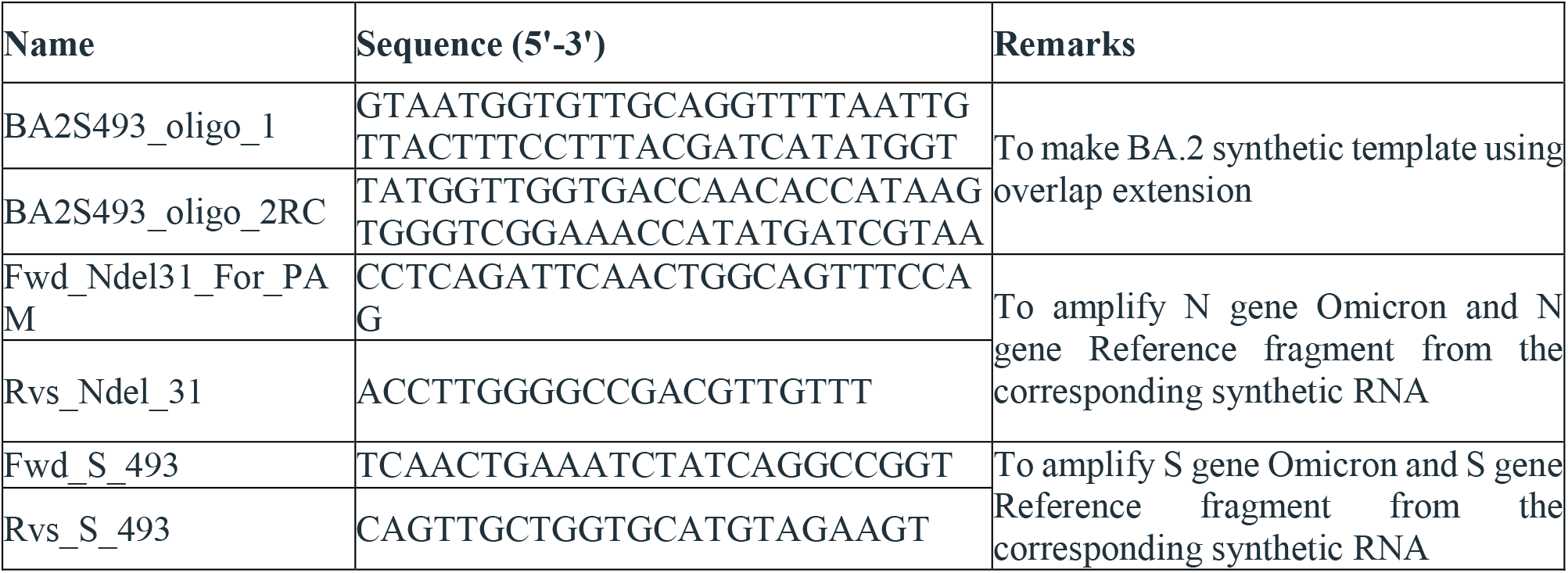
List of primes and oligonucleotides used for preparing the synthetic DNA templates.

**SI table 5:**
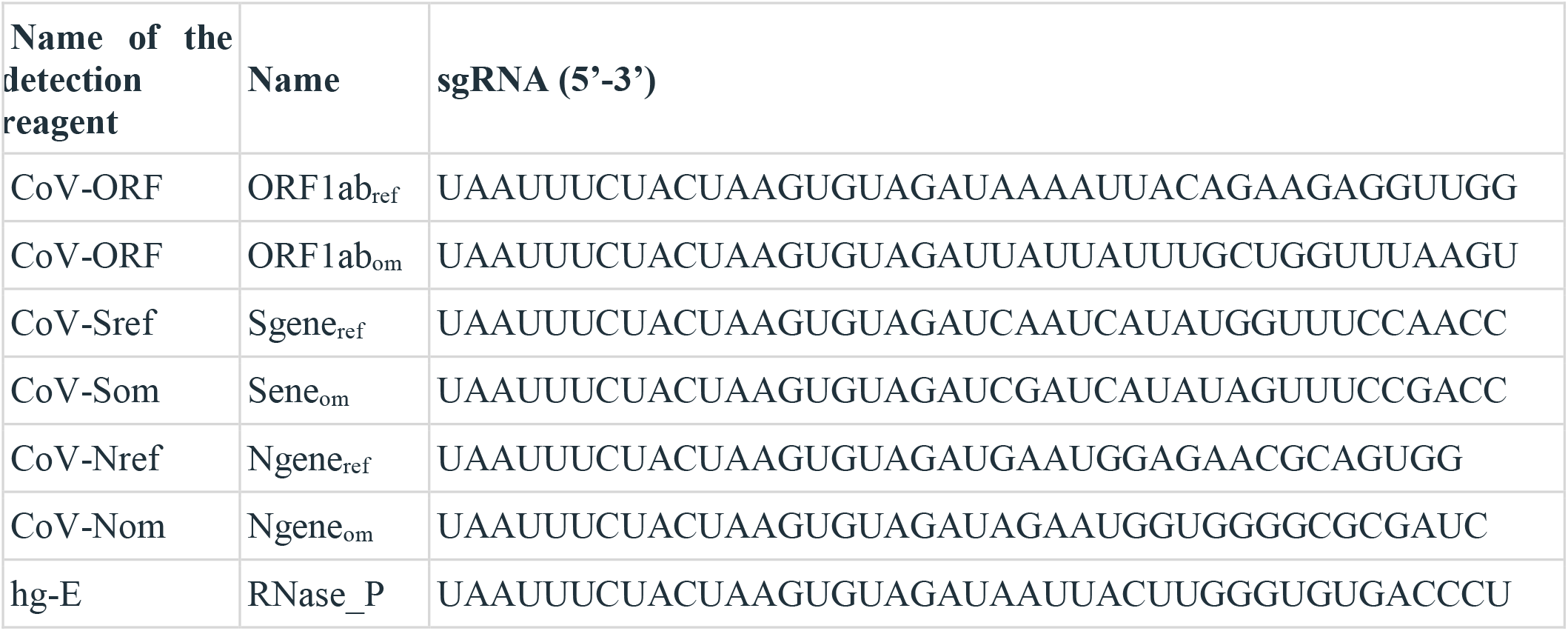
Sequence of the guide RNAs used in OmiCrisp

